# Comprehensive Blood Indicator PSI: A Novel Prognostic Tool for Resectable Colorectal Cancer

**DOI:** 10.1101/2023.07.28.23293251

**Authors:** Hao Cai, Jiancheng Li, Yu Chen, Qiao Zhang, Yang Liu, Houjun Jia

## Abstract

**Background:** Colorectal cancer (CRC) remains a major global health concern, with significant morbidity and mortality rates. Identifying reliable prognostic indicators is essential for optimizing risk stratification and guiding clinical management. In this study, we aimed to develop a comprehensive blood indicator based on systemic inflammation and nutritional condition to predict the prognosis of resectable CRC patients.

**Methods:** A retrospective cohort of 210 CRC patients who underwent radical resection at the First Affiliated Hospital of Chongqing Medical University, China, between January 2015 and December 2017, was included in the analysis. Baseline characteristics, preoperative blood markers, including neutrophil count, monocyte count, lymphocyte count, platelets, albumin, and CEA were retrospectively reviewed. Various blood indicators, such as NLR, PLR, MLR, SIRI and OPNI were calculated. The least absolute shrinkage and selection operator method (LASSO) was employed to select indicators to establish a novel comprehensive biomarker (named PSI). Kaplan-Meier survival curves and log-rank tests were used to evaluate the prognostic impact of preoperative OPNI, SIRI, and PSI. Univariate and multivariate Cox regression model were conducted to identify independent prognostic factors for CRC. The receiver operating characteristic (ROC) method assessed the predictive ability of PSI, stage, OPNI, and SIRI.

**Results:** Patients with higher preoperative OPNI and lower SIRI values had significantly better overall survival (OS). PSI was identified as an independent prognostic factor for OS in both univariate and multivariate analysis. Patients with medium (28.3-43.4) and high (>43.4) PSI scores exhibited superior OS compared to those with low (≤ 28.3) PSI scores. PSI showed higher predictive ability (AUC: 0.734) than individual indicators alone (OPNI: 0.721, SIRI: 0.645, stage: 0.635).

**Conclusion:** The novel comprehensive indicator, PSI, based on preoperative SIRI and OPNI, demonstrated significant prognostic value for resectable CRC patients. PSI outperformed individual indicators and could serve as a reliable tool for risk stratification and prognostic management in CRC patients.

## Introduction

Colorectal cancer (CRC) ranks as the third most prevalent malignant tumor and the second most fatal tumor-related disease globally, contributing to approximately 900,000 deaths annually [1,3]. Owing to the asymptomatic nature during early stages, 72% of CRC patients exhibit tumor cell invasion into adjacent tissues or lymph nodes at diagnosis, with 14% already presenting distant metastasis, significantly impacting overall prognosis [2]. Prognosis assessment in CRC traditionally relies on TNM staging classification; however, patients with CRC at the same pathological stage often experience varying prognosis, particularly in stages II and III [4-5]. Hence, a comprehensive evaluation of CRC prognosis is imperative, necessitating the development of robust prognostic indicators and identification of patients with adverse prognostic characteristics.

Tumor-associated inflammation is considered the 7th hallmark of cancer [6]. Early speculations on the relationship between tumor-associated inflammation and malignancy have led to a growing body of evidence supporting the role of inflammation in tumor microenvironment regulation, tumor cell proliferation, invasion, and immunosuppression [7,9]. Chronic inflammation plays a pivotal role in genomic destabilization, inducing DNA damage, affecting DNA repair systems, and altering cell cycle checkpoints, contributing to cellular carcinogenesis, including in colorectal cells [6]. The accumulation of mutations in genes such as TP53, within the context of chronic inflammation, initiates carcinogenesis in intestinal epithelial cells [11]. Subsequent genetic alterations, such as p53 gene deletions, promote the expression of inflammation-related genes, fostering a vicious cycle of inflammation promotion and malignant growth. For instance, functional p53 gene deletion increases nuclear factor-κB (NF-κB) transcription factor expression, contributing to CRC progression [10]. NF-κ B activation induces the expression of inflammatory cytokines, adhesion molecules, prostaglandin synthase pathway, nitric oxide synthase, and angiogenic factors, amplifying the systemic inflammatory response and promoting tumor angiogenesis, proliferation, immune evasion, and distant metastasis [6,8]. To predict patient survival, various blood markers, such as C-reactive protein [12], SIRI [13], SII [14], NLR [15], and PLR [15], have emerged as potential avenues in CRC prognostic management. However, these indicators predominantly focus on inflammation-related indicators and lack a comprehensive assessment of other significant prognostic factors.

Preoperative malnutrition is particularly prevalent in CRC patients and profoundly influences survival outcomes. Weight loss, muscle loss, and weakness resulting from decreased intake, metabolic changes, and gastrointestinal abnormalities contribute to adverse postoperative outcomes and long-term prognosis [16,17]. Remarkably, approximately 35% of CRC patients undergoing surgery suffer from preoperative moderate to severe malnutrition, a higher incidence than other non-gastrointestinal malignancies [16,18]. Preoperative malnutrition in CRC patients is associated with increased postoperative mortality within 30 days after surgery and elevated risks of complications, such as anastomotic fistulas, infections, septic shock, cardiovascular events, anastomotic leakage, and prolonged hospitalization [16,19,20]. Moreover, it predicts poorer overall and disease-free survival in the long term [16,21]. Onodera’s Prognostic Nutritional Index (OPNI), calculated as serum albumin + 5 × total lymphocyte count, has emerged as a common assessment tool for nutritional and immune status. Low preoperative OPNI values in CRC patients have been consistently associated with higher risks of serious postoperative complications and poorer overall survival.

In this study, we proposed a novel prognostic index integrating SIRI and OPNI to investigate its value in predicting long-term survival in resectable CRC cases. Our objective was to validate its predictive superiority and provide valuable research evidence for individualized prediction and decision-making in CRC management.

## Material and methods

### Patients

Between January 2015 and December 2017, a consecutive cohort of 260 patients with CRC who underwent radical resection at the First Affiliated Hospital of Chongqing Medical University (Chongqing, People’s Republic of China) were retrospectively reviewed. The exclusion criteria are as follows: 1) neoadjuvant radiotherapy or radiochemotherapy ahead of surgery; 2) patients with a history of other primary or secondary malignancies;3) patients who underwent emergency surgery prior to inclusion in the study; 4) patients with blood diseases, autoimmune disease or infection which may influence biomarkers. 5) patients who died directly or indirectly from diseases other than CRC. Finally, 210 cases were included based on the criteria above. The study has been approved by the independent Ethics Committee at The First Affiliated Hospital of Chongqing Medical University (K2023-304) and was performed following the ethical standards of the World Medical Association Declaration of Helsinki.

### Data collection

Patient baseline characteristics were retrospectively reviewed and collected through the electronic medical record system, such as age, gender, past medical history, family history, smoking history, and alcohol history. Blood markers were measured within three days before surgery and included absolute neutrophil count, monocyte count, absolute lymphocyte count, platelets, albumin, and tumor marker carcinoembryonic antigen (CEA). Then, some blood indicators were calculated accordingly: neutrophil × monocyte-to-lymphocyte ratio (SIRI,

Systemic Inflammation Response Index), serum albumin + 5 × total lymphocyte(OPNI, Onodera’s Prognostic Nutritional Index), neutrophil-to-lymphocyte ratio (NLR), platelet-to-lymphocyte ratio (PLR) and monocyte-to-lymphocyte ratio (MLR).

### Follow-up

All patients included in the study underwent radical resection. Patients with a high risk of local recurrence and distant metastasis were given adjuvant chemotherapy according to their wishes. Trained study interviewers followed up all patients via telephone. Overall survival (OS) was defined as the period from pathological diagnosis to death due to cancer or the most recent follow-up.

### Statistical analysis

The X-tile program determined the optimal cutoff values for NLR, PLR, MLR, SIRI, and OPNI (Yale University, Newhaven, Connecticut). Pearson’s χ2 test was applied to reveal the correlation between variables, and then the heat map showed the degree of correlation among the blood indicators. The LASSO algorithm was utilized to develop a novel and comprehensive indicator. The Kaplan-Meier method calculated survival curves and compared differences with the log-rank test. Hazard ratios (HRs) and 95% confidence intervals (CIs) were estimated based on the univariate and multivariate Cox regression model to find independent prognostic factors of CRC. The receiver operating characteristic (ROC) method was utilized to compare the predictive capability based on the area under the curve (AUC). A P-value less than 0.05 was statistical significance.

## Results

### Patients’ characteristics

Two hundred and ten patients were enrolled in the final analysis. The baseline characteristics are shown in **Table 1**. 118 (56.2%) were male patients, and 161 (78.5%) were diagnosed with stage II-III, which accounted for a large proportion of patients with outcome events. 75 (35.7%) patients had a BMI greater than 24 or less than 18.5, accounting for approximately half of the patients with a BMI within the 18.5-24 range.

**Table 1.**
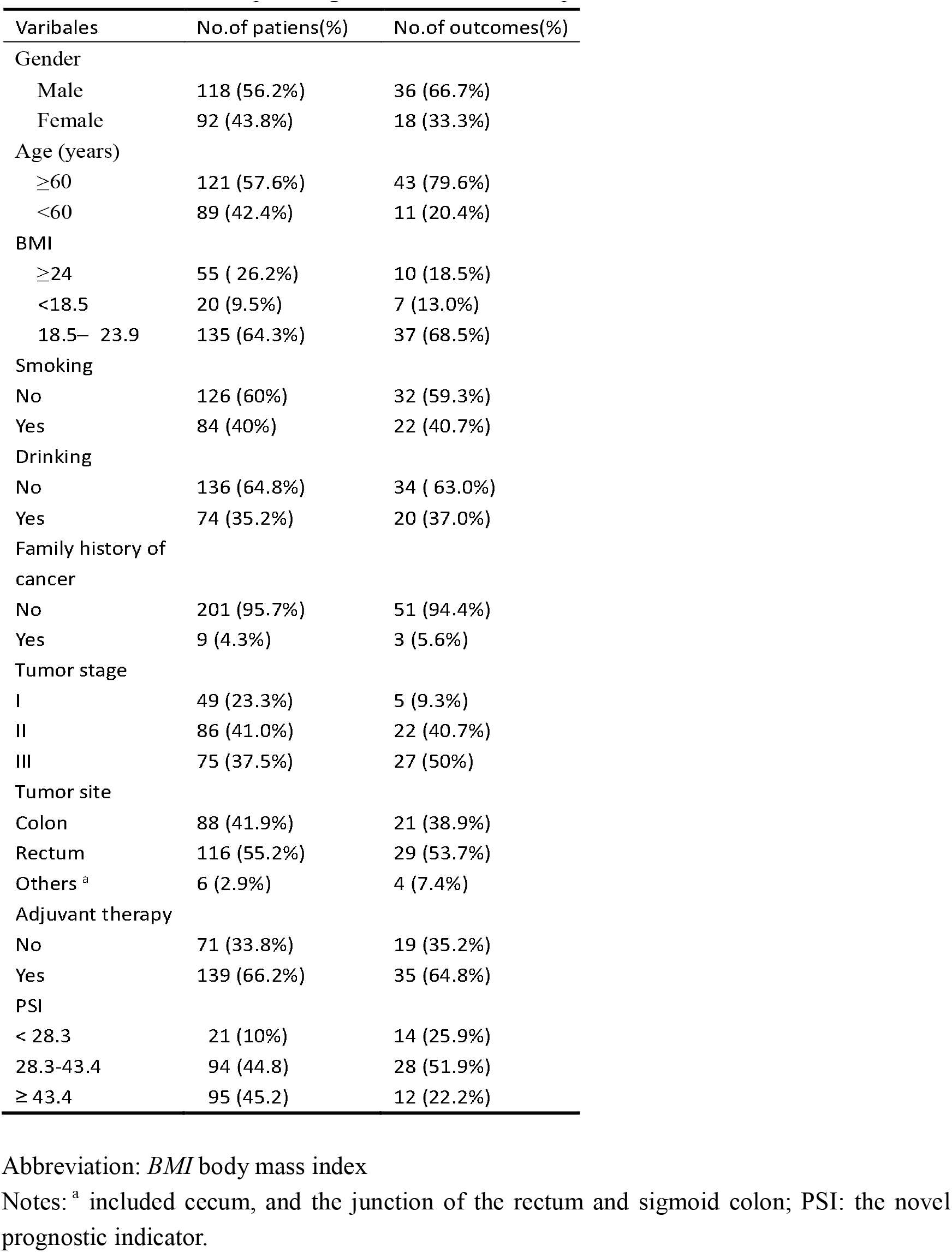
Baseline clinicopathological characteristics of patients with CRC.

| Varibales | No.of patiens(%) | No.of outcomes(%) |
| --- | --- | --- |
| Gender |  |  |
| Male | 118 (56.2%) | 36 (66.7%) |
| Female | 92 (43.8%) | 18 (33.3%) |
| Age (years) |  |  |
| ≥60 | 121 (57.6%) | 43 (79.6%) |
| <60 | 89 (42.4%) | 11 (20.4%) |
| BMI |  |  |
| ≥24 | 55 ( 26.2%) | 10 (18.5%) |
| <18.5 | 20 (9.5%) | 7 (13.0%) |
| 18.5– 23.9 | 135 (64.3%) | 37 (68.5%) |
| Smoking |  |  |
| No | 126 (60%) | 32 (59.3%) |
| Yes | 84 (40%) | 22 (40.7%) |
| Drinking |  |  |
| No | 136 (64.8%) | 34 ( 63.0%) |
| Yes | 74 (35.2%) | 20 (37.0%) |
| Family history of cancer |  |  |
| No | 201 (95.7%) | 51 (94.4%) |
| Yes | 9 (4.3%) | 3 (5.6%) |
| Tumor stage |  |  |
| I | 49 (23.3%) | 5 (9.3%) |
| II | 86 (41.0%) | 22 (40.7%) |
| III | 75 (37.5%) | 27 (50%) |
| Tumor site |  |  |
| Colon | 88 (41.9%) | 21 (38.9%) |
| Rectum | 116 (55.2%) | 29 (53.7%) |
| Others <sup>a</sup> | 6 (2.9%) | 4 (7.4%) |
| Adjuvant therapy |  |  |
| No | 71 (33.8%) | 19 (35.2%) |
| Yes | 139 (66.2%) | 35 (64.8%) |
| PSI |  |  |
| < 28.3 | 21 (10%) | 14 (25.9%) |
| 28.3-43.4 | 94 (44.8) | 28 (51.9%) |
| ≥ 43.4 | 95 (45.2) | 12 (22.2%) |

Abbreviation: *BMI* body mass index
Notes: <sup>a</sup> included cecum, and the junction of the rectum and sigmoid colon; PSI: the novel prognostic indicator.

### Correlation between preoperative blood indicators

The heat map depicted the correlations between the nine preoperative blood indicators, represented by the intensity of color shading (**Fig.1**). Strong correlations were observed among some indicators, with the MLR and MC displaying the strongest correlation. Pearson’s χ ^2^ test revealed that preoperative OPNI was significantly correlated with ALC, SII, SIRI, NLR, PLR, and MLR (p<0.001, p<0.001, p<0.01, p<0.001, p<0.001, p=0.001, respectively). However, no significant correlation was found between ANC and MC.

**Fig.1.**
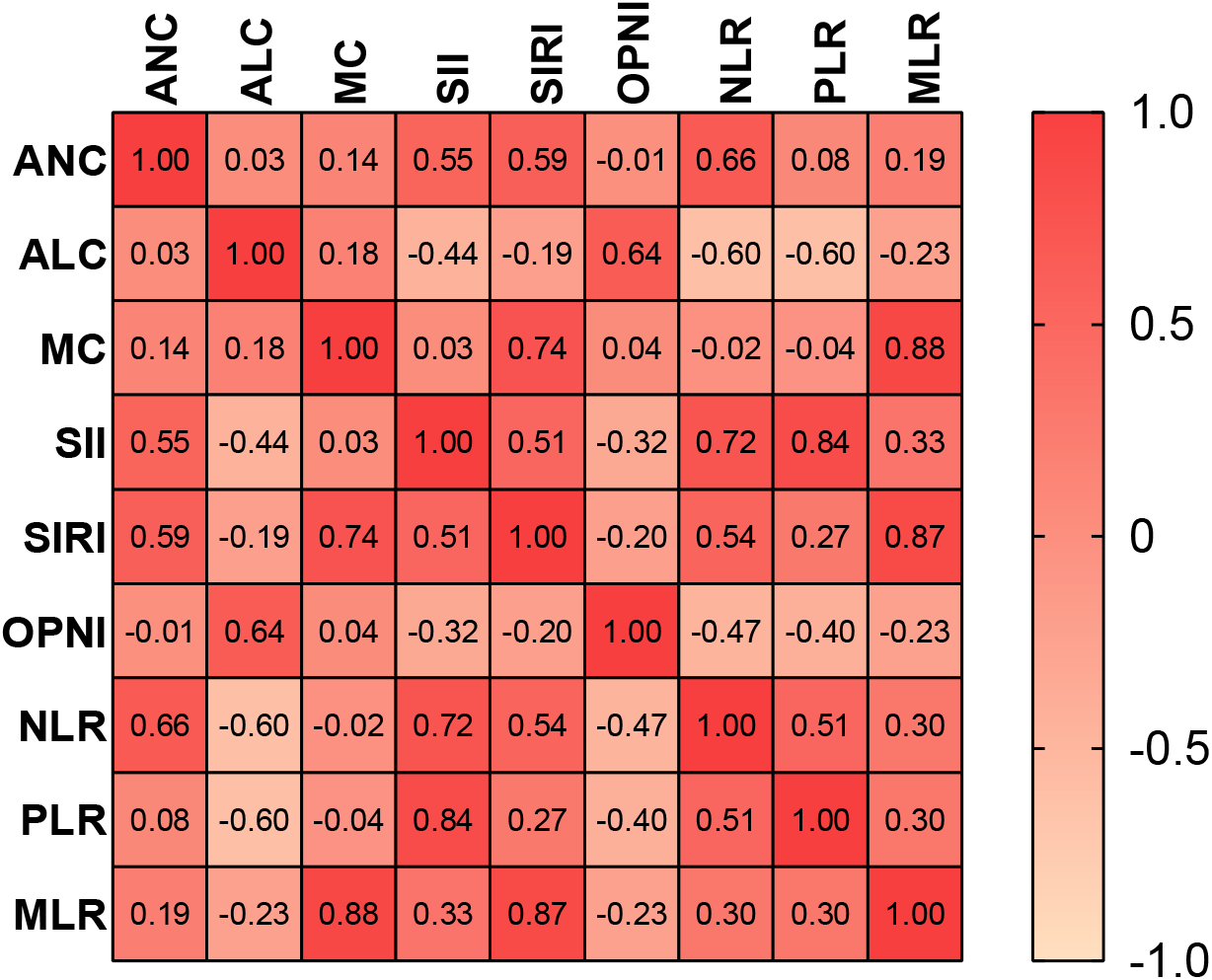
Correlation heat map of nine indicators. Abbreviations: *OPNI* Onodera’ s Prognostic Nutritional Index, *NLR* neutrophil-to-lymphocyte ratio, *PLR* platelet-to-lymphocyte ratio, *MLR* monocyte-to-lymphocyte ratio, *ANC* absolute neutrophil count, *ALC* absolute lymphocyte count, *MC* monocyte count, *SII s*ystemic immune-inflammation index

### Establishment of a comprehensive blood indicator

Considering the correlation between preoperative blood indicators and the limitations of traditional Cox regression modeling, the LASSO Cox regression model was employed to analyze the contribution of nine indicators to the endpoint event, assigning coefficients that help eliminate variables contributing minimally. As a result, the comprehensive indicator PSI was formulated as (0.9858×OPNI) - (7.2153×SIRI). Based on this formula, the 210 patients were categorized into three groups using the optimal cut-point obtained from X-tile software (Yale University, Newhaven, Connecticut): the high PSI group (≥ 43.4), the medium PSI group (28.3-43.4), and the low PSI group (< 28.3).

### Prognostic impact of preoperative OPNI or SIRI in resectable CRC

Patients with high levels of OPNI (≥ 49.6) exhibited significantly better overall survival (OS) compared to those with low OPNI values (<49.6) (p<0.001, **Fig.2B**). Similarly, patients with high SIRI values (≥ 2.0) demonstrated poorer survival in resectable CRC (p<0.001, **Fig.2A**).

**Fig.2.**
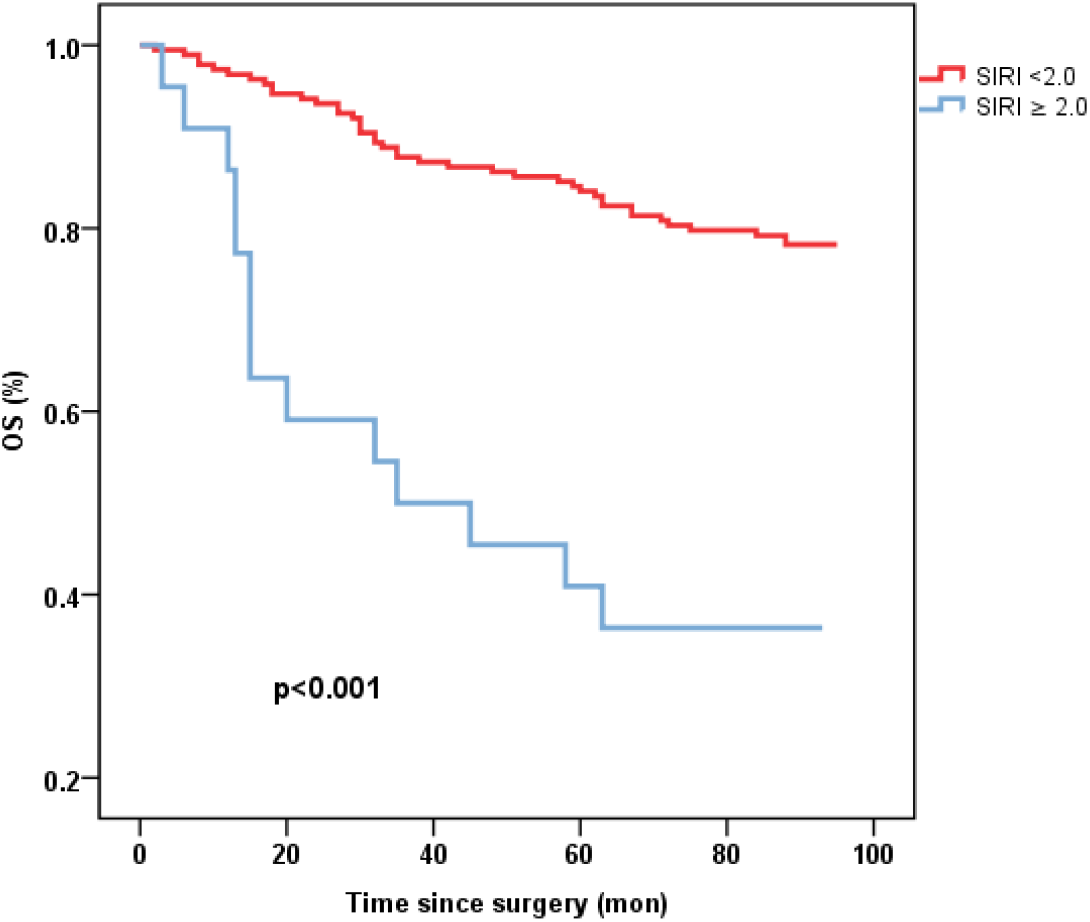
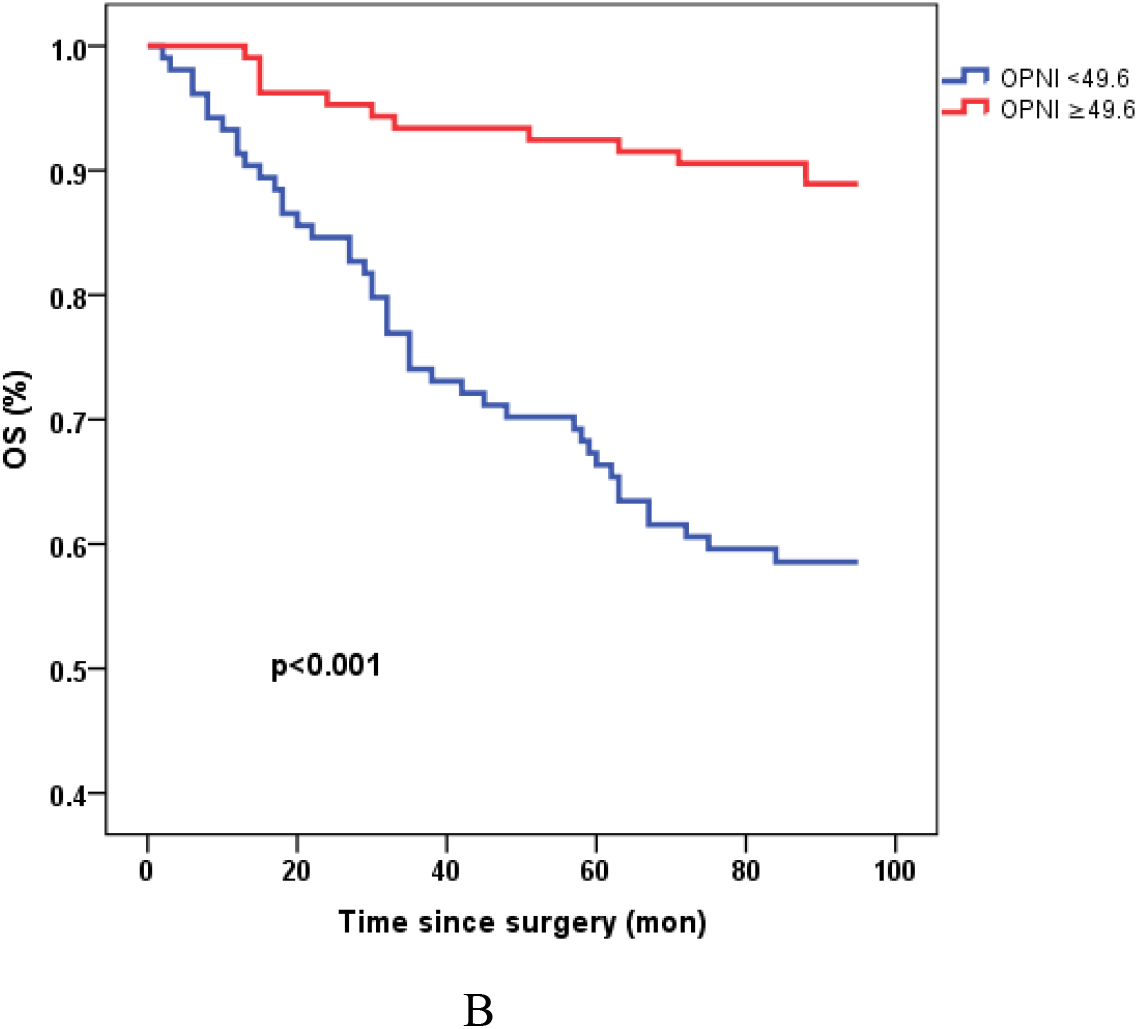
Kaplan-Meier survival curves of OS stratified by preoperative SIRI (**A**) and OPNI levels (**B**) in 210 resectable CRC patients (with log-rank test). Abbreviations: *OS* overall survival, *SIRI* systemic inflammation response index, *OPNI* Onodera’ s Prognostic Nutritional Index.

### Prognostic impact of preoperative PSI in resectable CRC

The prognostic value of PSI was assessed in comparison with lower counterparts, revealing that patients with higher PSI values (28.3-43.4 and >43.4) had notably improved OS (p<0.001, p<0.001, respectively, **Fig.3**). In both univariate and multivariate Cox models (**Table 2**), the PSI score was significantly associated with survival outcomes. Patients with a PSI score of 28.3-43.4 displayed a lower HR of 0.293 (95% CI: 0.154-0.558, p<0.001) and 0.312 (95% CI: 0.153-0.636, p=0.001) in univariate and multivariate Cox models, respectively. Likewise, for patients with a PSI score >43.4, the HR was 0.111 (95% CI: 0.051-0.242, p<0.001) in the univariate Cox model and 0.146 (95% CI: 0.060-0.351, p<0.001) in the multivariate Cox model, indicating a significantly reduced risk of poor survival. According to the results obtained from multivariate Cox regression analysis, PSI maintained its significance as an independent prognostic factor for OS (p≤0.001). Furthermore, PSI demonstrated superior predictive ability compared to stage, OPNI, and SIRI alone, with AUC values of 0.734 (95% CI: 0.654-0.815), 0.635 (95% CI: 0.552-0.717), 0.721 (95% CI: 0.642-0.800), and 0.645 (95% CI: 0.556-0.733), respectively (**Fig.4 A, B**).

**Fig.3.**
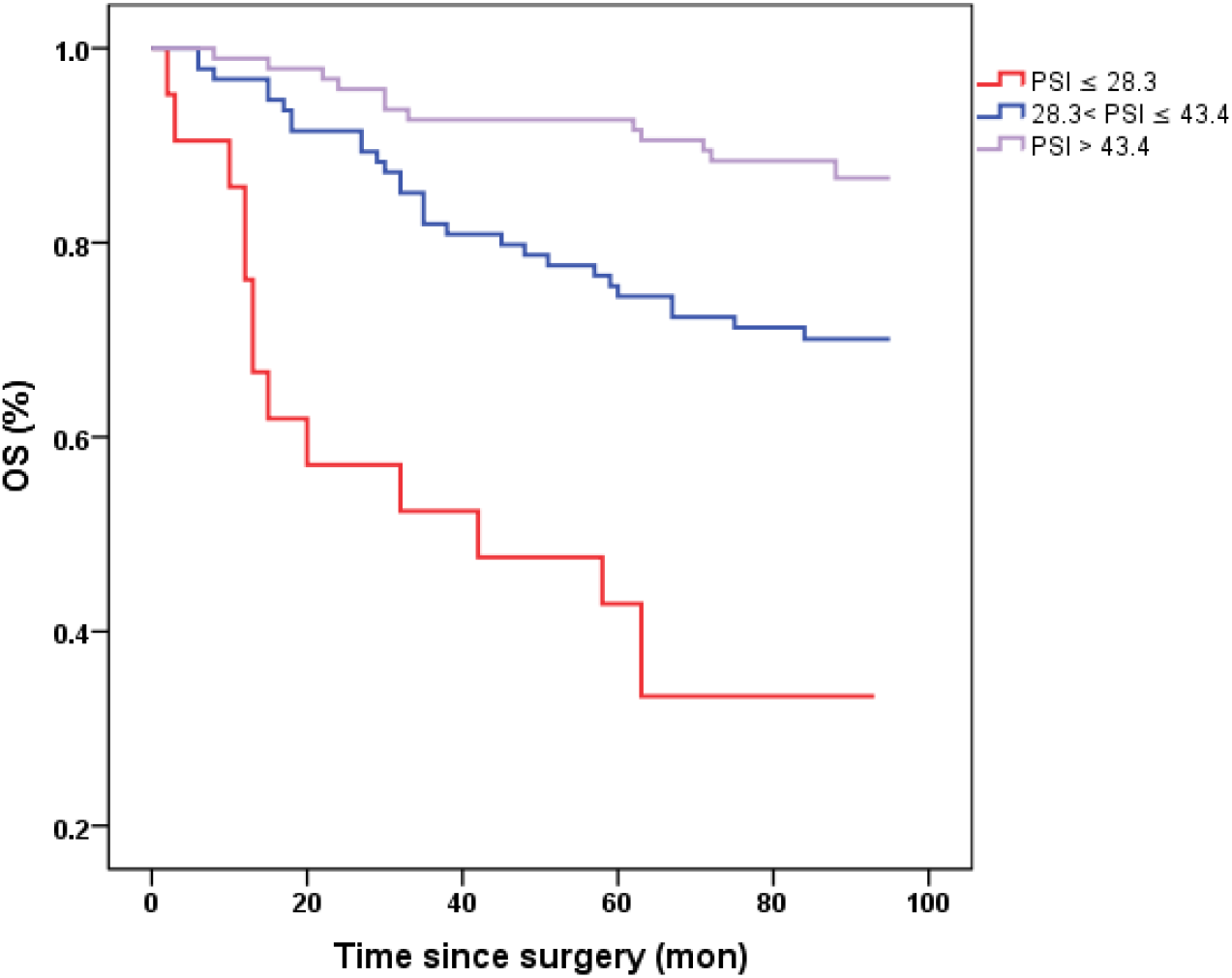
Kaplan-Meier survival curves of OS stratified by preoperative PSI in 210 resectable CRC patients (with log-rank test). Abbreviations: *OS* overall survival. Notes: the novel prognostic indicator (*PSI*).

**Table 2.**
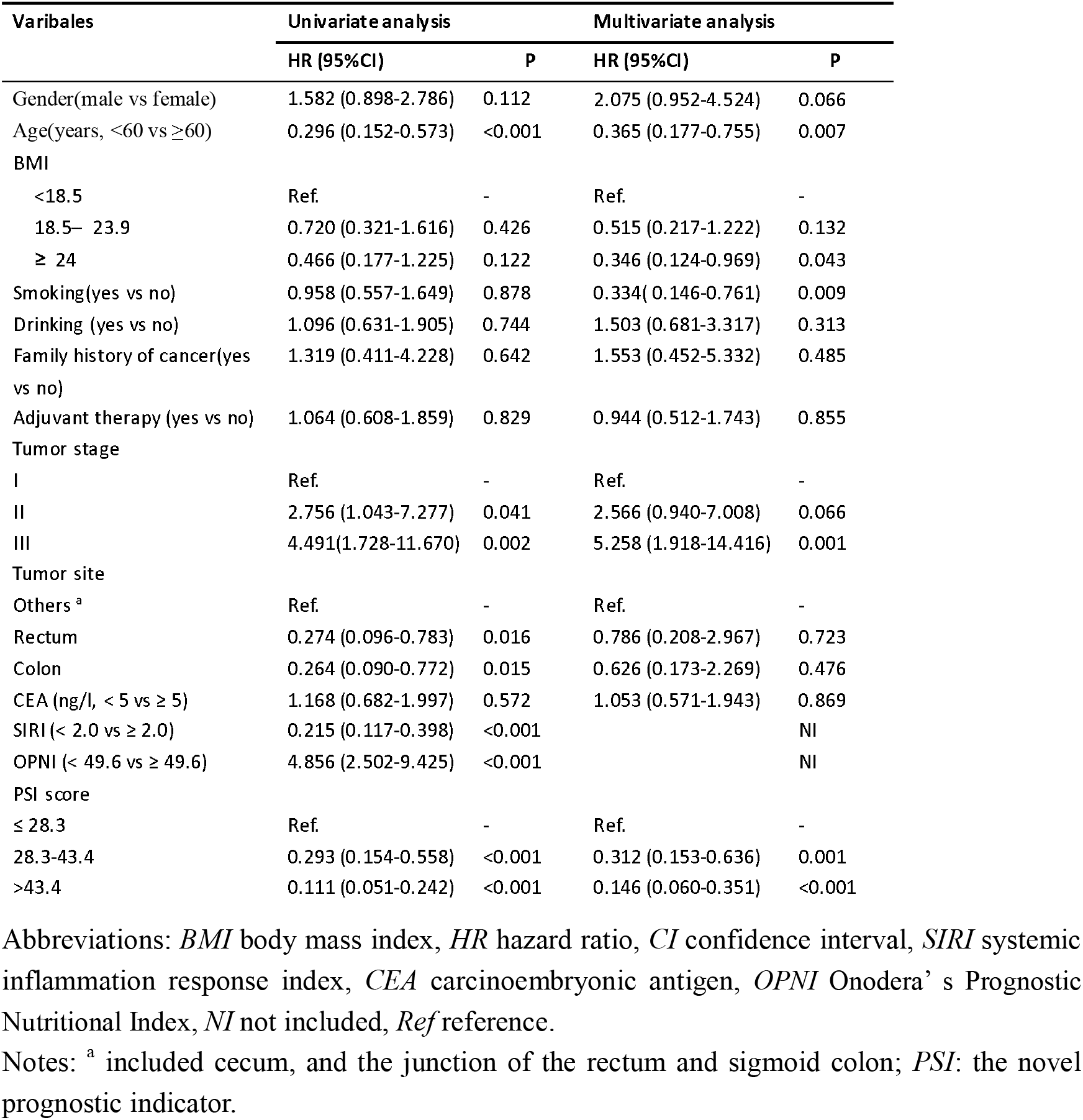
univariate and multivariate analysis for overall survival (OS)

| Variables | Univariate analysis |  | Multivariate analysis |  |
| --- | --- | --- | --- | --- |
|  | HR (95%CI) | P | HR (95%CI) | P |
| Gender(male vs female) | 1.582 (0.898-2.786) | 0.112 | 2.075 (0.952-4.524) | 0.066 |
| Age(years, <60 vs ≥60) | 0.296 (0.152-0.573) | <0.001 | 0.365 (0.177-0.755) | 0.007 |
| BMI |  |  |  |  |
| <18.5 | Ref. | - | Ref. | - |
| 18.5– 23.9 | 0.720 (0.321-1.616) | 0.426 | 0.515 (0.217-1.222) | 0.132 |
| ≥ 24 | 0.466 (0.177-1.225) | 0.122 | 0.346 (0.124-0.969) | 0.043 |
| Smoking(yes vs no) | 0.958 (0.557-1.649) | 0.878 | 0.334 (0.146-0.761) | 0.009 |
| Drinking (yes vs no) | 1.096 (0.631-1.905) | 0.744 | 1.503 (0.681-3.317) | 0.313 |
| Family history of cancer(yes vs no) | 1.319 (0.411-4.228) | 0.642 | 1.553 (0.452-5.332) | 0.485 |
| Adjuvant therapy (yes vs no) | 1.064 (0.608-1.859) | 0.829 | 0.944 (0.512-1.743) | 0.855 |
| Tumor stage |  |  |  |  |
| I | Ref. | - | Ref. | - |
| II | 2.756 (1.043-7.277) | 0.041 | 2.566 (0.940-7.008) | 0.066 |
| III | 4.491 (1.728-11.670) | 0.002 | 5.258 (1.918-14.416) | 0.001 |
| Tumor site |  |  |  |  |
| Others <sup>a</sup> | Ref. | - | Ref. | - |
| Rectum | 0.274 (0.096-0.783) | 0.016 | 0.786 (0.208-2.967) | 0.723 |
| Colon | 0.264 (0.090-0.772) | 0.015 | 0.626 (0.173-2.269) | 0.476 |
| CEA (ng/l, < 5 vs ≥ 5) | 1.168 (0.682-1.997) | 0.572 | 1.053 (0.571-1.943) | 0.869 |
| SIRI (< 2.0 vs ≥ 2.0) | 0.215 (0.117-0.398) | <0.001 |  | NI |
| OPNI (< 49.6 vs ≥ 49.6) | 4.856 (2.502-9.425) | <0.001 |  | NI |
| PSI score |  |  |  |  |
| ≤ 28.3 | Ref. | - | Ref. | - |
| 28.3-43.4 | 0.293 (0.154-0.558) | <0.001 | 0.312 (0.153-0.636) | 0.001 |
| >43.4 | 0.111 (0.051-0.242) | <0.001 | 0.146 (0.060-0.351) | <0.001 |

Abbreviations: *BMI* body mass index, *HR* hazard ratio, *CI* confidence interval, *SIRI* systemic inflammation response index, *CEA* carcinoembryonic antigen, *OPNI* Onodera's Prognostic Nutritional Index, *NI* not included, *Ref* reference.
Notes: <sup>a</sup> included cecum, and the junction of the rectum and sigmoid colon; *PSI*: the novel prognostic indicator.

**Fig.4.**
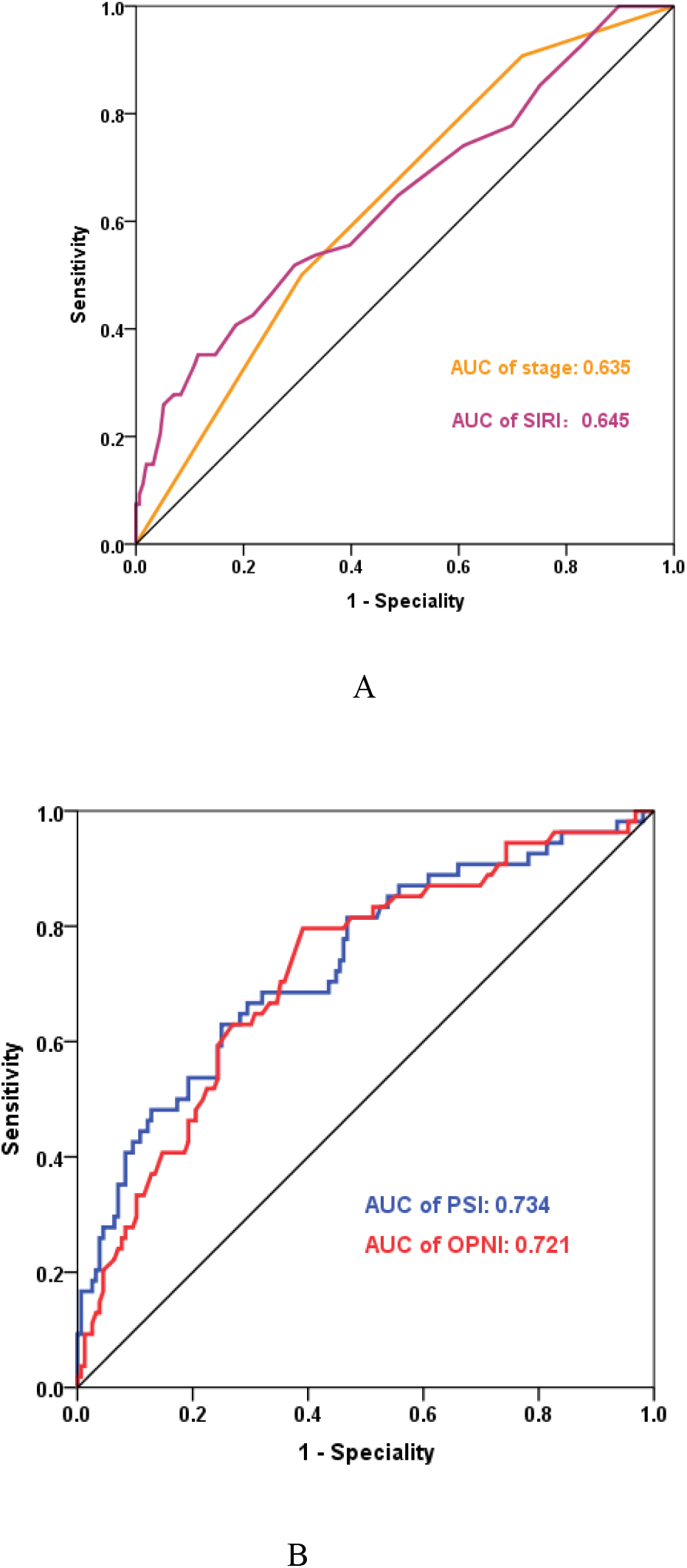
ROC curves for prognostic indicators in CRC - Categorized by Correlation (Positive or Negative). **A**: AUC of stage and SIRI; **B**: AUC of PSI and OPNI. Abbreviations: *AUC* Area Under the Curve, *OPNI* Onodera’ s Prognostic Nutritional Index, *SIRI* systemic inflammation response index. Notes: *PSI*: the novel prognostic indicator.

Additionally, age (<60 vs. ≥60) and tumor stage (stage III vs. I) were identified as independent prognostic factors. The HRs were 0.365 (95% CI: 0.177-0.755, p=0.007) and 5.258 (95% CI: 1.918-14.416, p=0.001), respectively. Moreover, site, SIRI, and OPNI were also found to have significant prognostic value in predicting the outcome of resectable CRC.

## Discussion

To the best of our knowledge, this study represents one of the few attempts to establish a comprehensive blood indicator for evaluating the prognosis of CRC patients by combining systemic inflammation and nutritional status. Our findings revealed that preoperative PSI served as an independent prognostic factor for CRC, with higher PSI levels associated with improved long-term survival. Moreover, the predictive value of PSI surpassed that of individual stage, OPNI, and SIRI in predicting clinical outcomes.

Systemic inflammation has been widely recognized as a critical factor promoting tumor proliferation, invasion, and metastasis, an established hallmark of malignancy [22,23]. Various blood markers have been explored to assess systemic inflammation and predict clinical outcomes in malignant diseases. Qi et al. were among the first to demonstrate that SIRI was associated with long-term survival in pancreatic cancer, serving as an independent prognostic factor for postoperative recurrence and clinical outcomes [35]. Subsequent studies validated preoperative SIRI levels as predictive of long-term survival in several malignancies, including gastric [24], lung [25], and breast cancer [26]. In the context of CRC, Cao et al. reported that higher preoperative SIRI levels were linked to poorer overall and disease-free survival [13]. Compared to other inflammation-related markers, such as NLR, PLR, and SII, SIRI demonstrated superior predictive value for CRC survival. Although evidence is accumulating, more research is still needed to fully understand the relationship between SIRI and CRC prognosis.

Preoperative malnutrition is more prevalent in CRC patients compared to other malignancies, often stemming from decreased food intake, intestinal obstruction, and abnormal gastrointestinal function caused by disease progression [27]. Numerous studies have confirmed its predictive ability for adverse short- and long-term outcomes in CRC, including surgical site infections, pneumonia, septic shock, and poorer survival [16,28,29]. Onodera and colleagues first proposed the Onodera’s Prognostic Nutritional Index (OPNI), which has since been widely adopted to assess preoperative nutritional status and predict prognosis in various malignancies, including oral cavity, colorectal, and liver cancers [30,31,32]. A high preoperative OPNI value was associated with better OS in CRC, establishing OPNI as an independent predictor for CRC cases [31]. Consequently, preoperative identification of malnourished patients allows for early initiation of nutritional support, forming the foundation for enhanced recovery after surgery (ERAS) protocols, reducing postoperative complications, and improving long-term prognosis [33,34].

While individual inflammatory or nutritional indicators exhibit prognostic predictive value, their accuracy and comprehensiveness can be enhanced. To address this, we utilized the LASSO method to analyze the contribution of nine indicators to the endpoint event, assigning coefficients that help eliminate variables contributing minimally. In this study, we established a comprehensive blood indicator (PSI) based on SIRI and OPNI and demonstrated that PSI more accurately predicted survival in resectable CRC patients. Our results indicated that both OPNI and SIRI were significant prognostic factors based on univariate analysis. Furthermore, multivariate analysis identified PSI as an independent prognostic predictor for overall survival (OS). Patients with PSI scores of 28.3-43.4 and >43.4 exhibited lower HRs of 0.312 (95% CI: 0.153-0.636, p=0.001) and 0.146 (95% CI: 0.060-0.351, p<0.001), respectively. Importantly, compared to single indicators, including stage, OPNI, and SIRI, PSI demonstrated superior prognostic survival prediction, with an AUC value of 0.734 (95% CI: 0.654-0.815). Additionally, our finding that high preoperative SIRI levels in CRC were associated with poorer OS aligns with the results of SIRI in prognostic studies of other malignancies. Thus, PSI stands as a reliable prognostic predictor, accurately assessing the systemic inflammation response and nutritional condition, thereby supporting a more comprehensive evaluation of CRC patient survival.

Nevertheless, this study does have certain limitations that warrant consideration. First, the included OPNI evaluated nutritional status at a single point in time, and subsequent nutritional improvements were not accounted for. Developing a dynamic evaluation system to assess patients’ nutritional status and consider its overall impact on short- and long-term prognosis may lead to a more comprehensive and accurate prognostic assessment. Second, the small sample size limits the generalizability of our results, necessitating external validation. Third, being a single-center study, potential selection bias may exist. Therefore, multi-center studies are still needed to corroborate our findings.

## Conclusion

In conclusion, our study presented a novel comprehensive blood indicator (PSI) based on SIRI and OPNI, demonstrating its potential as an independent prognostic factor for CRC. PSI outperformed individual inflammatory and nutritional markers in predicting clinical outcomes. This supports more accurate and comprehensive risk management and personalized treatment for resectable CRC patients.

## Data Availability

The datasets used in this study are available on request from the corresponding author.

## List of Abbreviations

CRC: colorectal cancer
OS: overall survival
DFS: disease free survival
SIRI: systemic inflammation response index
CEA: carcinoembryonic antigen
OPNI: Onodera’s prognostic nutritional Index
NLR: neutrophil-to-lymphocyte ratio
PLR: platelet-to-lymphocyte ratio
MLR: monocyte-to-lymphocyte ratio
BMI: body mass index
HR: hazard ratio
CI: confidence interval
NI: not included.
Ref: reference
SII: systemic immune-inflammation index
AUC: area under the curve
ANC: absolute neutrophil count
ALC: absolute lymphocyte count
MC: monocyte count

## Declarations

### Authors’ contribution

HC was in charge of acquiring and analyzing data, composing manuscripts, and conducting critical revisions. JCL took responsibility for patient follow-up and collecting clinicopathological data. YC, QZ, and YL provided technical support for data analysis and proofread the article for grammar. HJJ supervised the conception, design, and review of selected topics.

## Acknowledgement

None.

## Availability of data and materials

The datasets used in this study are available on request from the corresponding author.

## Ethics approval and consent to participate

This study was approved by the ethics committee of Chongqing Medical University(K2023-304) and followed the ethical standards of the Helsinki Declaration.

## Consent to participate

Not applicable.

